# COVID-19 mRNA Vaccination in Lactation: Assessment of adverse events and vaccine related antibodies in mother-infant dyads

**DOI:** 10.1101/2021.03.09.21253241

**Authors:** Yarden Golan, Mary Prahl, Arianna G. Cassidy, Caryl Gay, Alan H.B. Wu, Unurzul Jigmeddagva, Christine Y. Lin, Veronica J. Gonzalez, Emilia Basilio, Lakshmi Warrier, Sirirak Buarpung, Lin Li, Amy P. Murtha, Ifeyinwa V. Asiodu, Nadav Ahituv, Valerie J. Flaherman, Stephanie L. Gaw

**Affiliations:** Department of Bioengineering and Therapeutic Sciences, University of California, San Francisco, and Institute for Human Genetics, University of California, San Francisco, California, United States of America; Department of Pediatrics, University of California, San Francisco, California, United States of America; Division of Pediatric Infectious Diseases and Global Health, University of California, San Francisco, California, United States of America; Division of Maternal-Fetal Medicine, Department of Obstetrics, Gynecology, and Reproductive Sciences, University of California San Francisco, California, United States of America; Department of Family Health Care Nursing, University of California, San Francisco, California, United States of America; Department of Laboratory Medicine, University of California, San Francisco, California, United States of America; Center for Reproductive Sciences, Department of Obstetrics, Gynecology, and Reproductive Sciences, University of California San Francisco, California, United States of America; Department of Medicine, University of California, San Francisco, California, United States of America

**Keywords:** COVID-19, SARS-CoV-2, vaccine, lactation, immunity, antibodies, breastfeeding, human milk, mRNA vaccine, passive immunity, side effects

## Abstract

**Background:** Data regarding adverse events observed in the lactating mother-infant dyad and their immune response to COVID-19 mRNA vaccination during lactation are needed to inform vaccination guidelines.

**Methods:** From a prospective cohort of 50 lactating individuals who received mRNA-based vaccines for COVID-19 (mRNA-1273 and BNT162b2), blood and milk samples were collected prior to first vaccination dose, immediately prior to 2nd dose, and 4-10 weeks after 2nd dose. Symptoms in mother and infant were assessed by detailed questionnaires. Anti-SARS-CoV-2 antibody levels in blood and milk were measured by Pylon 3D automated immunoassay and ELISA. In addition, vaccine-related PEGylated proteins in milk were measured by ELISA. Blood samples were collected from a subset of infants whose mothers received the vaccine during lactation (4-15 weeks after mothers’ 2nd dose).

**Results:** No severe maternal or infant adverse events were reported in this cohort. Two mothers and two infants were diagnosed with COVID-19 during the study period. PEGylated proteins, were not found at significant levels in milk after vaccination. After vaccination, levels of anti-SARS-CoV-2 IgG and IgM significantly increased in maternal plasma and there was significant transfer of anti-SARS-CoV-2-Receptor Binding Domain (anti-RBD) IgA and IgG antibodies to milk. Milk IgA levels after the 2nd dose were negatively associated with infant age. Anti-SARS-CoV-2 IgG antibodies were not detected in the plasma of infants whose mothers were vaccinated during lactation.

**Conclusions:** COVID-19 mRNA vaccines generate robust immune responses in plasma and milk of lactating individuals without severe adverse events reported.

## Introduction

An important benefit of human milk is the presence of IgA and IgG antibodies that provide passive immunity to the infant (1,2). Anti-SARS-CoV-2 antibodies are present in milk from lactating women who were infected with SARS-CoV-2 (3,4) or who received COVID-19 mRNA vaccines (5–8). Specifically, high titers of anti-SARS-CoV-2 IgG were reported after vaccination (6). The function of these antibodies in protection of infants against COVID-19 is not fully understood. Recently, work has demonstrated that murine pups can transfer IgG antibodies from ingested milk to their bloodstream, via enteric Fc receptors (9); however, this phenomenon has not been demonstrated in humans. To better understand the function and distribution if human milk antibodies and infant immunization after maternal vaccination, we examined anti-SARS-CoV-2 antibody levels in infant blood and stool. In addition, information is lacking on the potential adverse effects of COVID-19 vaccination on lactating mothers and their infants, who were excluded from initial clinical trials of mRNA vaccination (10). Many breastfeeding individuals have concerns regarding the potential effects on their infant due to the lack of data, leading to delayed vaccination or early cessation of breastfeeding (11).

In this study, we examined blood and milk samples from lactating mothers who received a COVID-19 mRNA vaccine and their infants for the presence of anti-SARS-CoV2 antibodies, presence of PEGylated protein vaccine products, and self-reported vaccine-related symptoms in order to address the gap of knowledge regarding vaccination efficacy and safety in this population.

## Methods

### Study approval and study population

The institutional review board of the University of California San Francisco approved the study. Written, informed consent was obtained from all study volunteers in the COVID-19 Vaccine in Pregnancy and Lactation (COVIPAL) cohort study from December 2020 to June 2021. Eligible participants were actively lactating, planning to receive any COVID-19 vaccine, and willing to donate blood and/or milk samples.

### Clinical data collection

Clinical data on vaccine side effects were collected through an online questionnaire that was sent to participants 21 days or more after each vaccine administration. Questionnaires were distributed using REDCap.

### Sample collection

Maternal blood and milk samples were collected at three time points: 1) up to 1 day before the 1st dose (pre-vaccine); 2) on the day of and prior to administration of the 2nd dose (after 1st dose); and between 4-10 weeks after the 2nd dose (after 2nd dose). In some cases, additional milk samples were collected up to 31 days before the 1st dose, 24 hours after each dose, and weekly for up to 4 weeks after the 2nd dose. Infant blood was collected by heel stick by trained study staff at 5-15 weeks after 2nd maternal vaccination.

### Milk processing

Fresh human milk samples were self-collected by participants into sterile containers at several time points before, during, and after vaccination. Milk samples were either collected immediately by the study staff or frozen by mothers in their home freezer as soon as possible after pumping. Samples were kept on ice during transport from home to the lab for processing. Milk was aliquoted and stored in -80°C until analyzed.

### Measurement of SARS-CoV-2 specific IgM and IgG in plasma samples

Whole blood was collected into tubes containing EDTA. Plasma was isolated from whole blood by centrifugation and immediately cryopreserved at -80°C until analysis. Anti-SARS-CoV-2 plasma IgM and IgG antibodies were measured using the Pylon 3D automated immunoassay system(12) (ET Healthcare, Palo Alto, CA). In brief, quartz glass probes pre-coated with either affinity-purified goat anti-human IgM (IgM capture) or Protein G (IgG capture) were dipped into diluted plasma samples, washed, and then dipped into the assay reagent containing both biotinylated, recombinant spike protein receptor binding domain (RBD) and nucleocapsid protein (NP). After washing, the probes were incubated with Cy®5-streptavidin (Cy5-SA) polysaccharide conjugate reagent, allowing for cyclic amplification of the fluorescence signal. The background-corrected signal of SARS-CoV-2 specific antibodies was reported as relative fluorescent units (RFU). IgM and IgG measurements greater than 50 RFU were considered positive RFUs.

### Measurement of IgA and IgG by ELISA assay in milk

After thawing, milk fat was separated by cold centrifugation (10,000g for 10 min, 4°C). Milk supernatant samples were diluted 1:2 in sample diluent buffer and were plated in duplicate on a 96-well plate containing S1 spike protein RBD (Ray-Biotech, GA, USA, IEQ-CoVS1RBD-IgG-1 and IEQ-CoVS1RBD-IgA-1). For monomeric IgA assays, samples were also plated in duplicate on a second 96-well plate coated with human albumin to account for non-specific binding. OD values for albumin were subtracted from the OD values for RBD. Each plate contained seven wells of serial dilutions (1:3) of a positive control from an inactivated serum sample which contains SARS-COV-2 S1 RBD protein human IgA antibody (provided with the kit) and one blank negative control. The mean absorbance of each sample was captured on an ELISA plate reader at 450 nm. Background values (blank negative control) were subtracted from the albumin and RBD plates. Standard controls were used to create a standard curve and determine the level of anti-RBD IgA and IgG in unit/ml.

### Measurement of Polyethylene Glycol (PEGylated) proteins in human milk by ELISA

Milk supernatant was diluted 1:8 with the provided sample buffer and analyzed by PEGylated protein ELISA kit (Enzo, Farmingdale, NY, USA). Seven wells of each plate were loaded with serial dilutions (1:2) of mRNA-1273 or BNT162b2 to generate the standard curve for each vaccine (**Figure S1**). In addition, vaccines were inoculated into human milk samples at three different concentrations (33µl/ml, 3.3µl/ml and 0.33µl/ml) and were analyzed separately to ensure the ability of the kit to detect the vaccine PEGylated components in milk samples (**Figure S1**). Prism 9 (v 9.1.2) was used to interpolate PEGylated proteins concentration in the samples based on OD values, using a sigmoidal, four parameters logistic curve. Standard curve for mRNA-1273 or BNT162b2 were used to analyse participant milk samples based on the vaccine received. Of note, the assay measures all types of PEGylated proteins (if present in the sample), and not only the vaccine PEGylated proteins.

### Statistics

All data analyses were conducted using Stata statistical software (v14, College Station, TX). Descriptive statistics included frequencies for categorical variables, and means, standard deviations, medians, and ranges for continuous variables. Group differences in categorical variables were analyzed using Fisher’s exact test, and group differences in continuous variables were analyzed using Mann-Whiney U tests. McNemar tests were used to evaluate differences in symptom frequencies after each vaccine dose. Spearman correlations was used to assess the magnitude of associations between continuous variables. Non-parametric tests were used to accommodate non-normal distributions and small group sizes.

## Results

### Participant characteristics

During the study period, 50 participants answered all study questionnaires, provided blood and/or milk samples, had an infant up to 18 months old and were included in this analysis. Two infants were diagnosed with COVID-19 during this study (**Table S1**, infant of participants 1 and 2). One mother reported that her infant had mild symptoms 1 week after the 2nd dose; this infant’s vaccinated mother had a negative test at the time of the infant’s positive PCR test. A second infant had positive plasma anti-SARS-CoV-2 IgG and IgA, despite the mother receiving the vaccine postpartum and reported no known prior COVID-19 infection. The mother’s plasma was subsequently found to be positive to antibodies against SARS-CoV-2 nucleocapsid protein, indicating a likely natural asymptomatic SARS-CoV-2 infection (further details in **Table S1**). Two mothers were positive for COVID-19 and are presented in **Table S1** (participants 2 and 3); they were excluded from further analysis of symptomatology. Cohort characteristics are presented in **Table 1**. Twenty-seven female participants (mean age 35.7 years (± 3.9)) received the BNT162b2 vaccine (Pfizer, 56%), and 21 received the mRNA-1237 (Moderna, 44%). The mean infant age at mother’s 1st dose was 5 months (± 3.9). All mothers continued to feed their infants with milk at the time of the 2nd vaccination, and all except one continued up to the time of follow up sample collection (4-10 weeks after 2nd dose). There were no significant differences in maternal or infant characteristics by vaccine manufacturer.

**Table 1.** Sample characteristics overall and by vaccine manufacturer.

| Sample Characteristics | Full Cohort<br>(n=48, 100%) | BNT162b2<br>(n=27, 56%) | mRNA-1237<br>(n=21, 44%) |
| --- | --- | --- | --- |
| ***Maternal characteristics*** |  |  |  |
| Maternal age, years |  |  |  |
| Median (min, max) | 35 (27, 46) | 35 (30, 45) | 35 (27, 46) |
| Race/ethnicity, % (n) |  |  |  |
| Asian | 31% (15) | 30% (8) | 33% (7) |
| Black or African American | 2% (1) | 4% (1) | 0% (0) |
| White/Caucasian | 59% (28) | 55% (15) | 62% (13) |
| Other (Arab) | 2% (1) | 0% (0) | 5% (1) |
| More than 1 race/ethnicity<br>(White+Latina/Asian/Middle Eastern) | 6% (3) | 11% (3) | 0% (0) |
| Highest level of education completed |  |  |  |
| Some college | 2% (1) | 0% (0) | 5% (1) |
| College graduate | 17% (8) | 19% (5) | 14% (3) |
| Advanced degree | 81% (39) | 81% (22) | 81% (17) |
| Work in health care? |  |  |  |
| Yes, providing direct patient care | 58% (28) | 52% (14) | 67% (14) |
| Yes, but not in direct patient care | 19% (9) | 15% (4) | 24% (5) |
| No | 23% (11) | 33% (9) | 9% (2) |
| Pre-Pregnancy Body Mass Index |  |  |  |
| Median (min, max) | 23.4 (19.1,<br>37.5) | 23.4 (19.1,<br>35.9) | 22.8 (19.6,<br>37.5) |
| Number of children |  |  |  |
| 1 | 40% (19) | 41% (11) | 38% (8) |
| 2 | 46% (22) | 41% (11) | 52% (11) |
| 3 | 12% (6) | 15% (4) | 10% (2) |
| 4 | 2% (1) | 3% (1) | 0% (0) |
| Duration of most recent pregnancy,<br>weeks |  |  |  |
| Median (min, max) | 39.0 (33.9,<br>41.1) | 39.1 (33.9,<br>41.0) | 39.0 (37.4,<br>41.1) |
| ***Infant characteristics*** |  |  |  |
| Infant age at maternal 1st dose,<br>months |  |  |  |
| Median (min, max) | 4.7 (0.1,<br>17.2) | 4.8 (0.2,<br>15.2) | 4.6 (0.1,<br>17.2) |
| Sex, % (n) |  |  |  |
| Male | 60% (29) | 67% (18) | 52% (11) |
| Female | 40% (19) | 33% (9) | 48% (10) |
| Exclusively breastfeeding (and no solids) |  |  |  |
| Yes | 23% (11) | 22% (6) | 24% (5) |
| No | 77% (37) | 78% (21) | 76% (16) |
| ***Days after vaccine that symptoms were assessed*** |  |  |  |
| Dose 1 |  |  |  |
| Mean (SD) | 78.7 (31.8) | 78.3 (35.4) | 79.2 (27.4) |
| Median (min, max) | 81 (18, 154] | 78 (18, 154) | 86 (26, 117) |
| Dose 2 |  |  |  |
| Mean (SD) | 59.6 (25.1) | 62.6 (28.3) | 55.8 (20.2) |
| Median (min, max) | 58.5 (28, 133) | 57 (29, 133) | 60 (28, 89) |

Note: None of the characteristics above differed significantly by vaccine manufacturer.
Abbreviations: Standard deviation (SD), minimum (min), maximum (max)

### Post vaccination symptoms

Self-reported symptoms after each vaccine dose are presented in **Table 2**. Fever, chills, headache, joint pain, muscle aches or body aches, and fatigue or tiredness were reported by significantly more participants after the 2nd dose than after the 1st dose (**Table 2**). All 21 participants (100%) who received the mRNA-1237 vaccine reported injection site symptoms, while only 21 (78%) of 27 BNT-162b2 recipients reported injection site symptoms (p=0.02) (**Table 2**). Two mothers reported slightly less milk production in the first 24-72 hours after vaccine doses (**Table 2**). With respect to infant symptoms, 12% of mothers reported at least one symptom after the 1st maternal vaccine dose (primarily gastrointestinal symptoms and sleep changes), and none reported an infant symptom after the 2nd dose (**Table 3**). In summary, no severe adverse events (death, life threatening, hospitalization, disability) for mothers or nursing infants were reported in this cohort after vaccination, and reported symptoms resolved up to 72 hours after vaccination.

**Table 2:** Symptoms after each vaccine dose.

| Symptoms | Full Cohort: |  |  | After 1st dose |  |  | After 2nd dose |  |  |
| --- | --- | --- | --- | --- | --- | --- | --- | --- | --- |
|  | 1st dose | 2nd dose | P-value<sup>a</sup> | BNT162b2 | mRNA-1237 | p-value<sup>b</sup> | BNT162b2 | mRNA-1237 | p-value<sup>b</sup> |
|  | n=48 |  |  | n=27 | n=21 |  | n=27 | n=21 |  |
| Injection site symptoms, % (n) |  |  |  |  |  |  |  |  |  |
| Any injection site symptoms | 88% (42) | 88% (42) | >0.99 | 78% (21) | 100% (21) | **0.02** | 78% (21) | 100% (21) | **.02** |
| Pain | 88% (42) | 85% (41) | 0.71 | 78% (21) | 100% (21) | **0.02** | 78% (21) | 95% (20) | 0.12 |
| Redness | 4% (2) | 10% (5) | 0.08 | 0% (0) | 10% (2) | 0.19 | 4% (1) | 19% (4) | 0.15 |
| Swelling | 17% (8) | 17% (8) | >0.99 | 7% (2) | 29% (6) | 0.12 | 11% (3) | 24% (5) | 0.27 |
| Itching | 4% (2) | 4% (2) | >0.99 | 4% (1) | 5% (1) | >.99 | 4% (1) | 5% (1) | >.99 |
| Rash around injection site | 2% (1) | 4% (2) | 0.32 | 0% (0) | 5% (1) | 0.44 | 0% (0) | 10% (2) | 0.19 |
| Generalized symptoms, % (n) |  |  |  |  |  |  |  |  |  |
| Any general symptoms | 48% (23) | 92% (44) | **<0.001** | 44% (12) | 52% (11) | 0.77 | 85% (23) | 100% (21) | 0.12 |
| Fever | 12% (6) | 62% (30) | **<0.001** | 19% (5) | 5% (1) | 0.21 | 52% (14) | 76% (16) | 0.13 |
| Chills | 8% (4) | 48% (23) | **<0.001** | 11% (3) | 5% (1) | 0.62 | 37% (10) | 62% (13) | 0.14 |
| Headache | 21% (10) | 67% (32) | **<0.001** | 11% (3) | 33% (7) | 0.08 | 56% (15) | 81% (17) | 0.07 |
| Joint pain | 8% (4) | 31% (15) | **0.002** | 7% (2) | 10% (2) | >0.99 | 30% (8) | 33% (7) | >0.99 |
| Muscle/body aches | 21% (10) | 69% (33) | **<0.001** | 30% (8) | 10% (2) | 0.15 | 59% (16) | 81% (17) | 0.13 |
| Fatigue or tiredness | 44%<br>(21) | 81%<br>(39) | **<0.001** | 41% (11) | 48%<br>(10) | 0.77 | 67% (18) | 100%<br>(21) | **0.003** |
| Nausea | 4%<br>(2) | 12%<br>(6) | 0.10 | 4% (1) | 5% (1) | >0.99 | 7% (2) | 19% (4) | 0.38 |
| Vomiting | 0%<br>(0) | 0%<br>(0) | --- | 0% (0) | 0% (0) | --- | 0% (0) | 0% (0) | ---- |
| Diarrhea | 4%<br>(2) | 4%<br>(2) | >0.99 | 4% (1) | 5% (1) | >0.99 | 0% (0) | 10% (2) | 0.19 |
| Abdominal pain | 2%<br>(1) | 0%<br>(0) | 0.32 | 0% (0) | 5% (1) | 0.44 | 0% (0) | 0% (0) | ---- |
| Rash not near injection site | 0%<br>(0) | 0%<br>(0) | --- | 0% (0) | 0% (0) | --- | 0% (0) | 0% (0) | ---- |
| Lump/swelling in breast (same side as injection) | 0%<br>(0) | 2%<br>(1) | 0.32 | 0% (0) | 0% (0) | --- | 4% (1) | 0% (0) | >0.99 |
| Lump/swelling in breast (opposite side as injection) | 0%<br>(0) | 0%<br>(0) | --- | 0% (0) | 0% (0) | --- | 0% (0) | 0% (0) | ---- |
| Mastitis | 2%<br>(1) | 0%<br>(0) | 0.32 | 0% (0) | 5% (1) | 0.44 | 0% (0) | 0% (0) | ---- |
| Decrease in milk supply | 2%<br>(1) | 2%<br>(1) | >0.99 | 0% (0) | 5% (1) | 0.44 | 0% (0) | 5% (1) | 0.44 |

<sup>a</sup> McNemar's test; <sup>b</sup> Fisher's Exact test

**Table 3.**
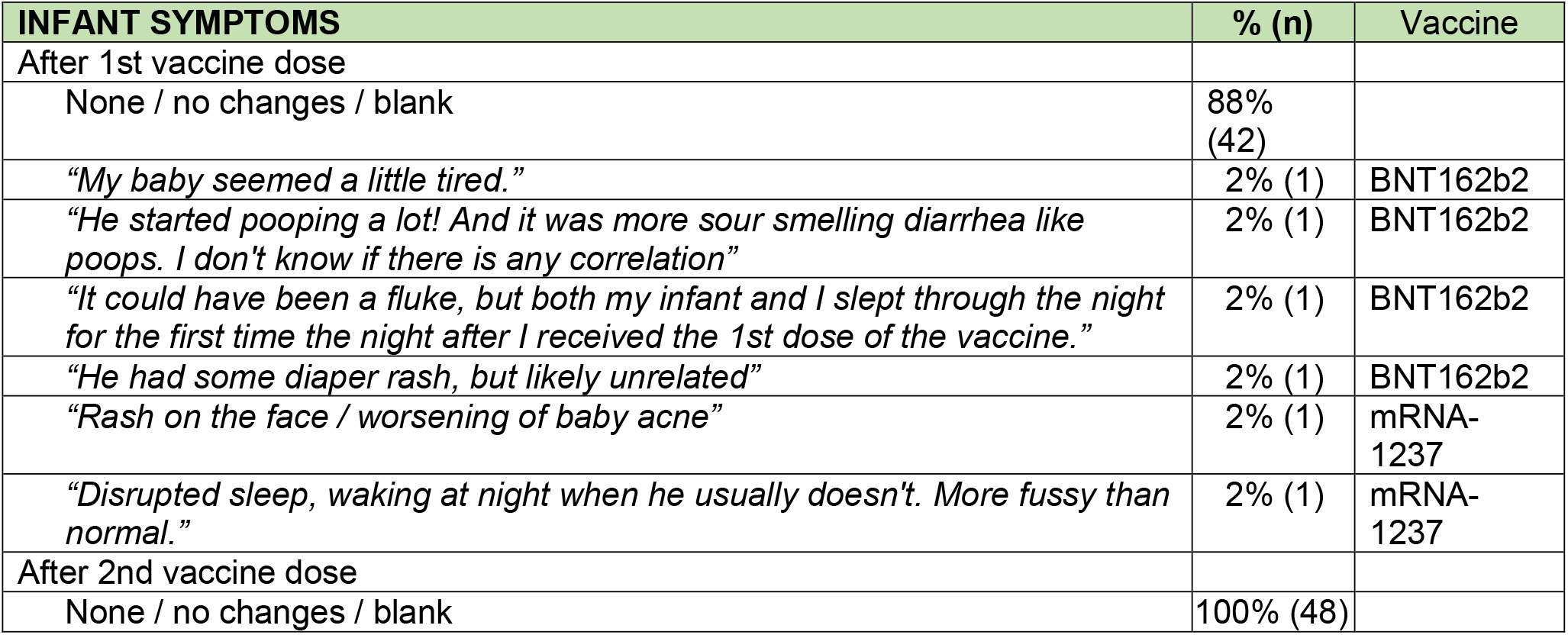
Infant symptoms reported after maternal vaccination (write-in only)

### PEG detection in human milk

Polyethylene glycol (PEG) is present in the lipid nanoparticles of the mRNA-based vaccines, and was reported to cause allergic reaction after vaccination in rare cases (13,14). To address concerns about vaccine components passing to milk after vaccination, we performed ELISA assays to measure PEG-ylated proteins levels in milk after vaccination from 13 participants. PEG-ylated proteins were measured in milk samples collected before the vaccine, and at various time points post-vaccination (from 24 hours after 1st dose to 2 weeks after 2nd dose). Pre-vaccine PEG-ylated proteins concentration did not significantly differ from PEG-ylated proteins levels at any post-vaccine time point in either paired or unpaired comparisons (**Figure 1**).

**Figure 1:**
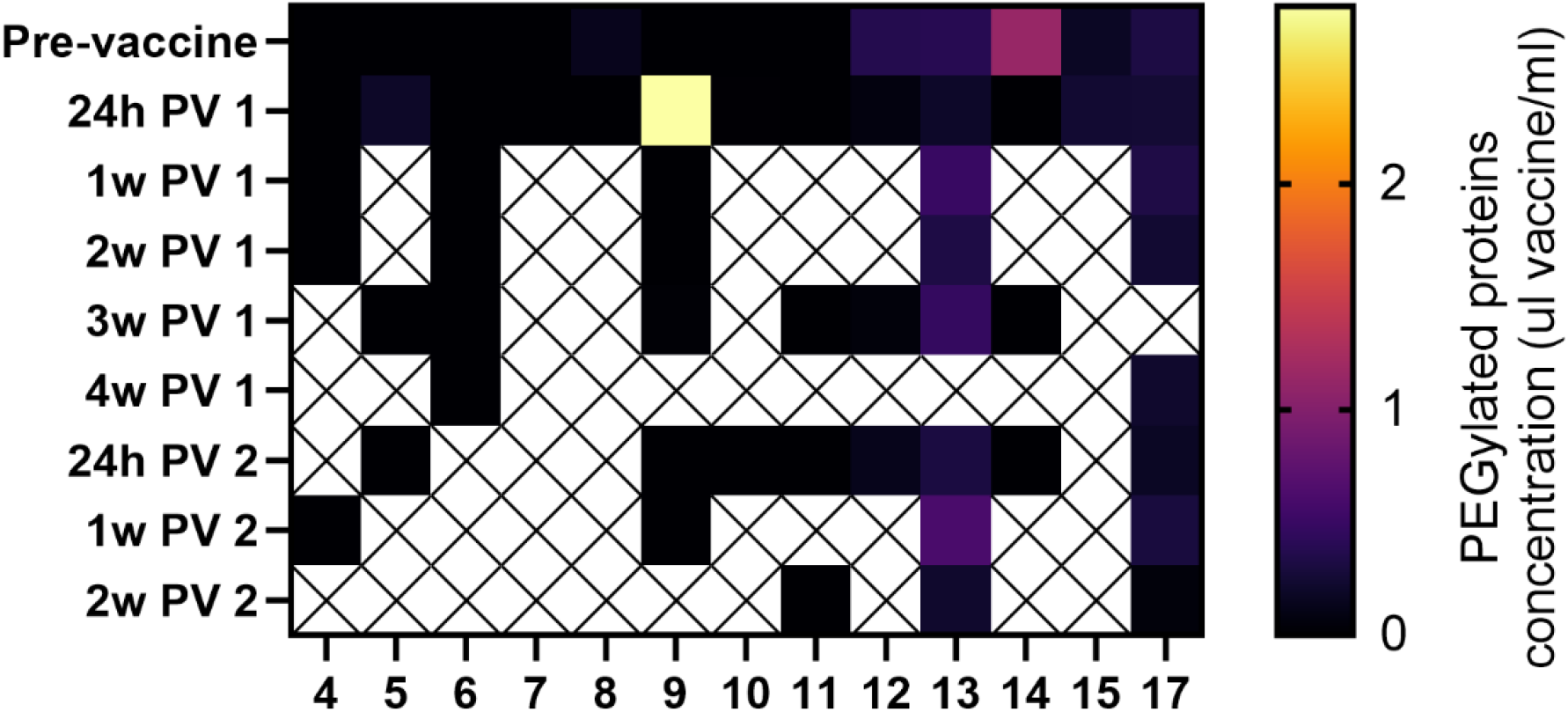
Detection of vaccine PEG in human milk samples. PEGylated protein concentration in each sample were interpolated based on vaccine standard curves (Figure S1). No significant differences were observed between samples collected at any of the post vaccine (PV) time points and the pre-vaccine samples (paired and unpaired two-tailed t-tests). Y axes represent time of sample collection, as hours (h) or weeks (w) Post vaccine 1 (PV 1), or Post vaccine 2 (PV 2).

### Anti-SARS-CoV-2 antibody levels in blood and milk samples after vaccination

We analyzed blood and milk samples from lactating individuals for anti-SARS-CoV-2 antibodies to measure immune response after vaccination. Maternal blood anti-SARS-CoV-2 IgM and IgG antibodies increased significantly after the 1st dose **(Figure 2)**. Anti-SARS-CoV-2 IgM levels were not significantly higher 4-10 weeks after the 2nd dose compared to samples collected after dose 1 (on the day of the 2nd dose) (**Figure 2A and 2B)**. In contrast, anti-SARS-CoV-2 IgG levels increased significantly after the 2nd dose (P value <0.0001) when compared to samples collected immediately prior to the 2nd dose (**Figure 2C and 2D)**. There was no significant difference in blood antibody levels between participants who received the mRNA-1237 compared to the BNT-162b2 vaccine after dose 2 (determined by unpaired Mann-Whitney test).

**Figure 2:**
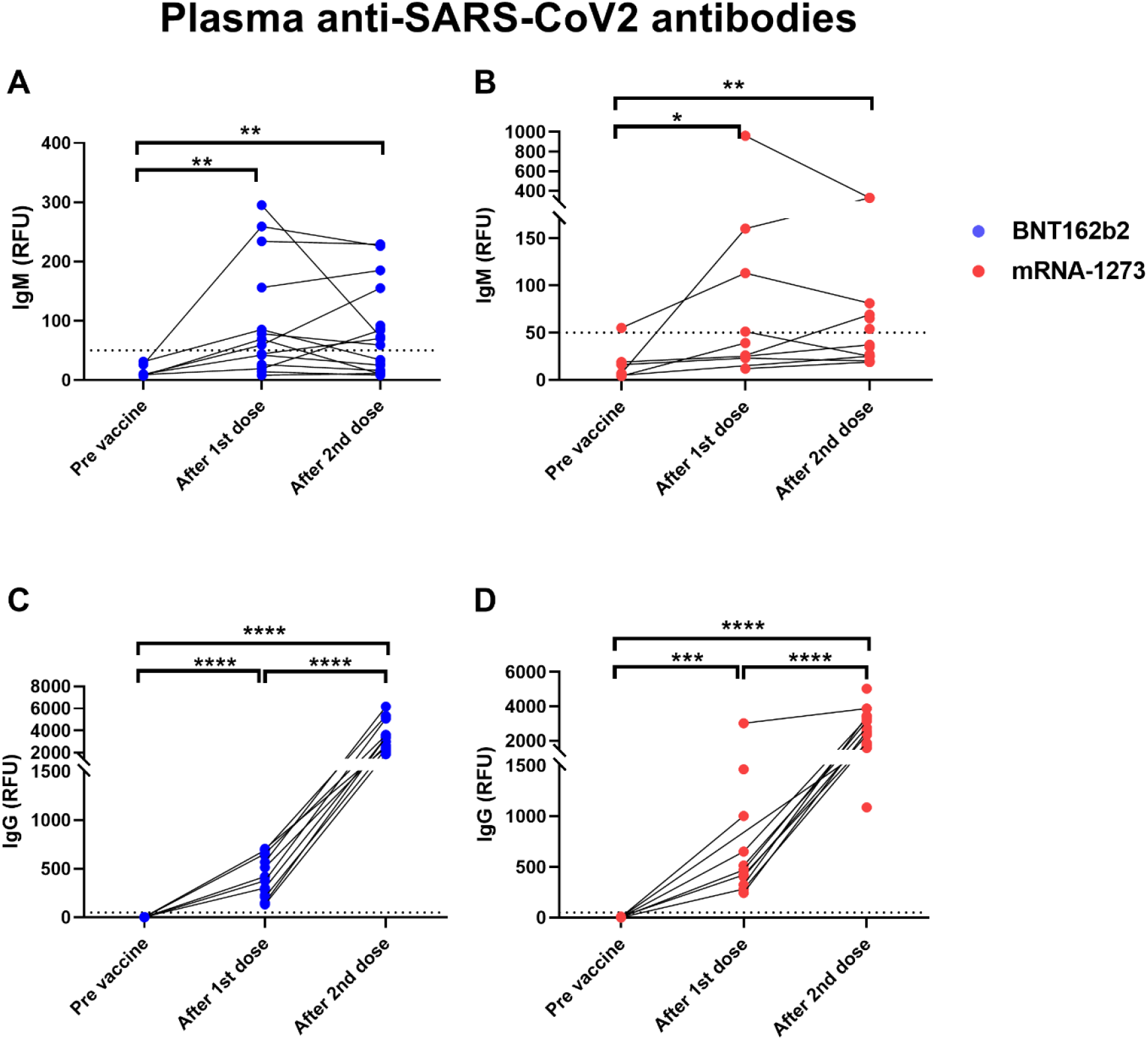
Elevated levels of plasma anti-SARS-CoV2 antibodies in COVID-19 mRNA vaccinated lactating individuals. Anti-SARS-CoV2 IgM levels in plasma of lactating individuals receiving BNT-162b2 (n=19) (A) and mRNA-1273 (n=13) (B) COVID-19 vaccines (RFU-relative fluorescent units, dashed line represents positive cut-off >50 RFU). Anti-SARS-CoV2 IgG levels in plasma of lactating individuals receiving BNT-162b2 (C) and mRNA-1273 (D) COVID-19 vaccines. After 1st dose samples were collected on the day of the second vaccine, and after 2nd dose samples were collect 4-10 weeks post 2nd dose. Asterisks represent p-values: *= p-value <0.05, **= p-value <0.01, ***= <0.001, ****= <0.0001 as determined by unpaired Mann-Whitney test.

We found significantly higher levels of IgA antibodies specific to SARS-CoV-2 RBD protein in human milk samples collected after the 1st dose of both BNT-162b2 and mRNA-1237 vaccines (**Figure 3A and 3B**). There was no significant increase in milk anti-RBD IgA after the 2nd vaccination as compared to after dose 1 (**Figure 3A and 3B**). Twelve individuals (25%, BNT-162b2 n=7; mRNA-1237 n=5) did not have detectable levels of anti-RBD IgA after either the 1st or 2nd dose (infants age at 1st dose range 1-11 months). Milk anti-RBD IgG levels increased after the 1st dose of vaccine and increased further after the 2nd dose (**Figure 3C and 3D**). There were no significant differences in milk anti-RBD IgG levels between women who received BNT-162b2 (**Figure 3C)** and mRNA-1237 (**Figure 3D)**. These findings suggest that mRNA vaccine results in a robust immune response leading to increased anti SARS-CoV-2 antibody levels in blood, but also in milk during lactation.

**Figure 3:**
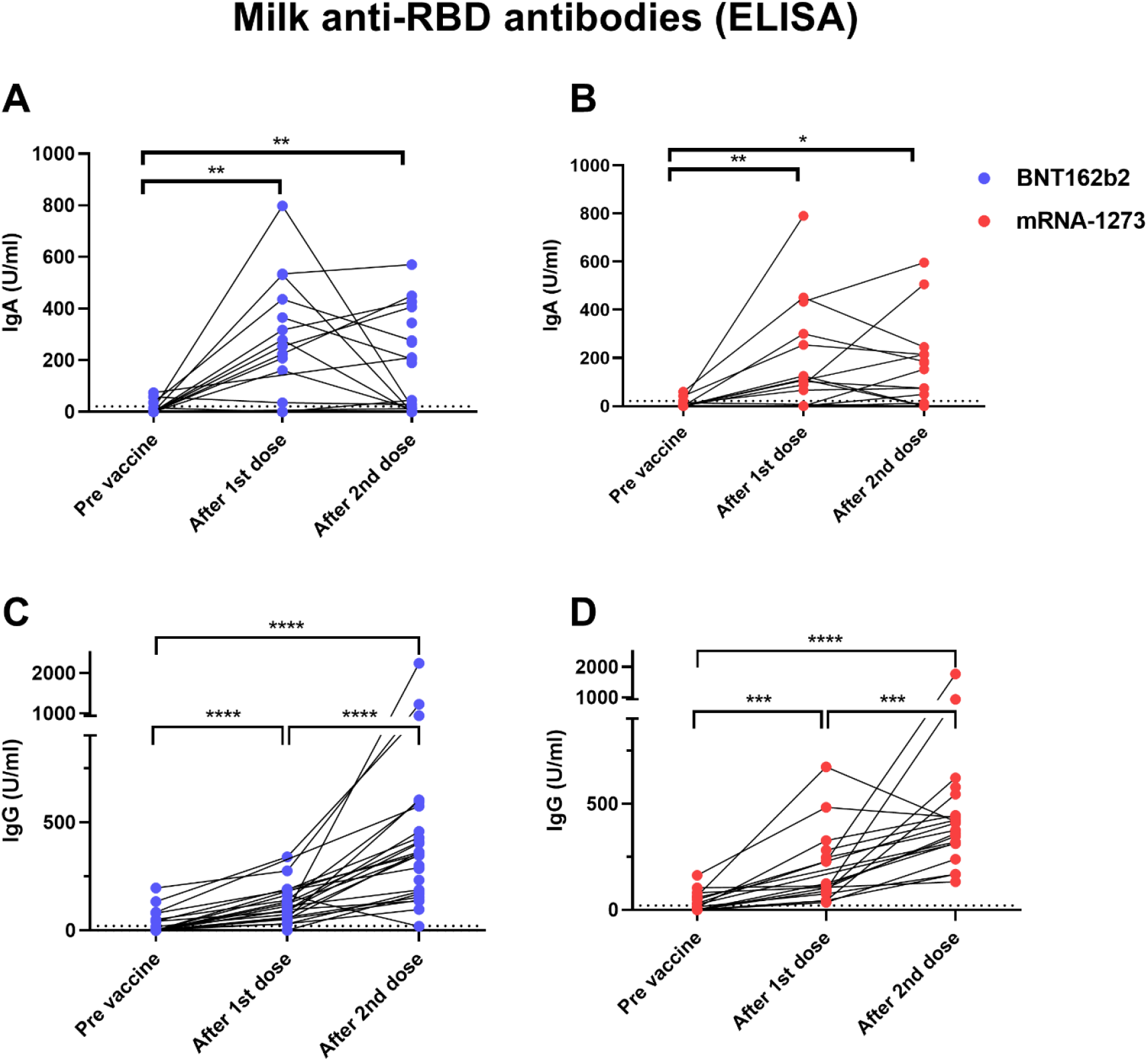
Elevated levels of milk anti-SARS-CoV2 IgA antibodies in COVID-19 mRNA vaccinated lactating individuals. Milk samples from individuals receiving BNT-162b2 (n=27) (A) and mRNA-1273 (n=21) (B) COVID-19 vaccines were analyzed for anti-SARS-CoV2 IgA antibodies using ELISA at various time points as indicated on the X axis. After 1st dose samples were collected on the day of the second vaccine, and after 2nd dose samples were collect 4-10 weeks post 2nd dose. Milk anti-SARS-CoV2 IgG levels were measured using ELISA in milk samples from individuals receiving BNT-162b2 (n=27) (C) or mRNA-1273 (n=21) (D). Asterisks represent p-values: *= p-value <0.05, **= p-value <0.01, ***= <0.001 as determined by unpaired Mann-Whitney test. Dashed line represents positive cut-off >21 U/ml.

### Correlations between antibody levels, participant characteristics, and symptoms

In order to better understand the differences in antibody responses between individuals in our cohort, we performed multiple correlation tests to determine whether IgG and IgA antibodies levels correlated with timing of sample collection after vaccination (range 4-10 weeks after 2nd dose), infant age at time of vaccination, or maternal BMI (**Table S2)**. Milk IgA (but not IgG) levels declined significantly as the infant age at time of vaccination increased **(Figure 4A and 4B)**. There was no significant correlation between IgG and IgA levels and either the length of time after 2nd dose or maternal BMI (**Table S2 and S3**). The levels of IgG and IgA antibodies induced in milk were significantly correlated after 1st dose (**Figure 4C)**, but not after the 2nd dose (**Figure 4D**). There was no correlation between the anti SARS-CoV-2 IgG levels in blood and milk after 1st dose (**Figure 4E**), but there was a positive correlation between levels at 4-10 weeks after 2nd dose (**Figure 4F**).

**Figure 4:**
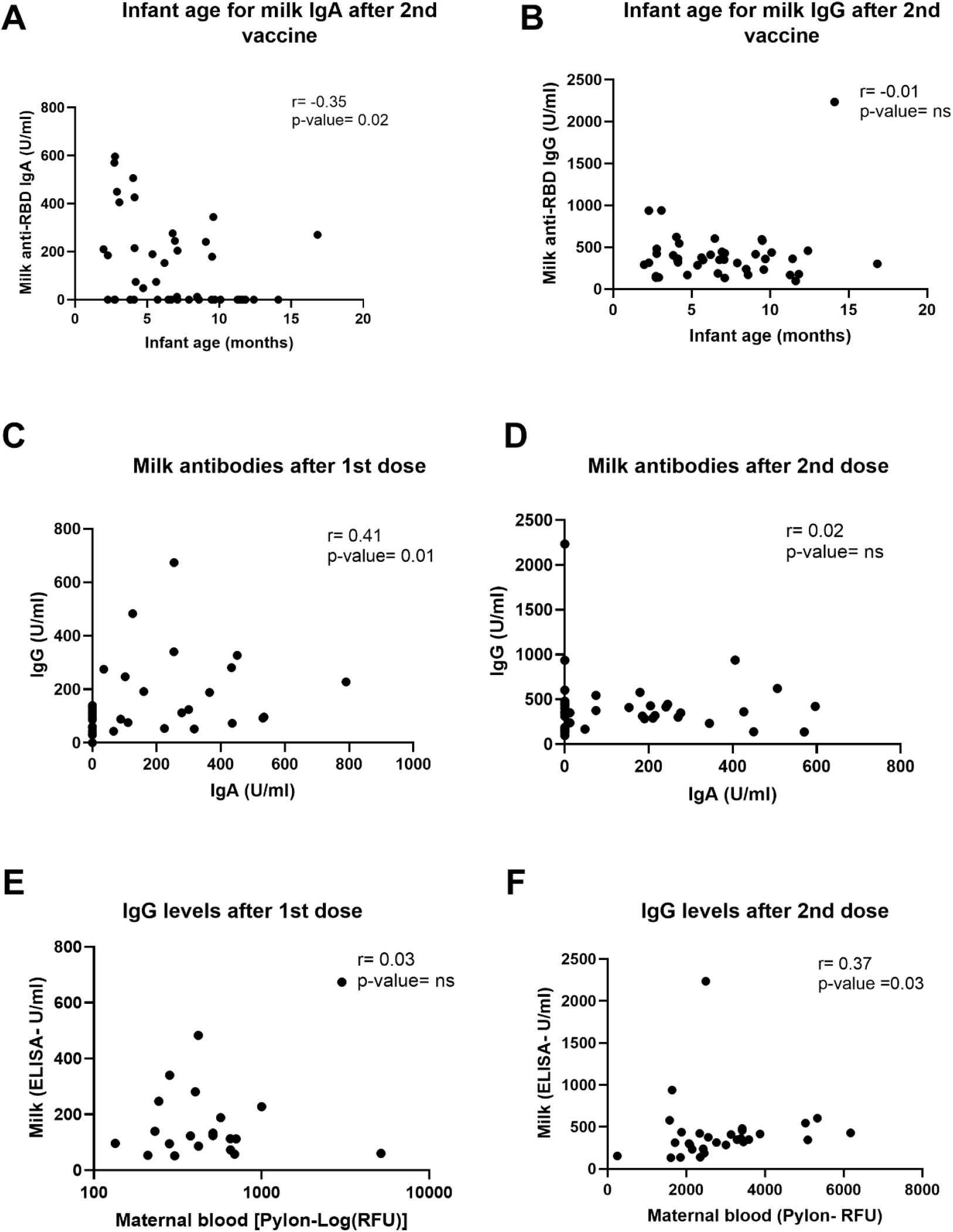
Correlations between milk antibodies, blood antibodies and infant age. Two-tailed Spearman correlation was used to correlate milk IgA (A) and IgG (B) levels (Y axis) and infant age (X axis) 4-10 weeks after the 2nd dose administration (n=30). In addition, two-tailed Spearman correlation was used to correlate milk IgG (Y axis) and milk IgA levels (X axis) on the day of 2nd dose administration(C), 21-28 days after 1st dose (n=35) and and 4-10 weeks after 2nd dose (D). We also tested correlation between milk (Y axis) and maternal plasma (X axis) IgG levels at day of 2nd dose (E) and 4-10 weeks after the 2nd dose administration (F) (n=30). Semi-partial correlations were used to assess relationships between variables while controlling for the effects of other relevant variables.

### Plasma levels of anti-SARS-CoV-2 IgG are not detectable in infants after maternal vaccination during lactation

Although maternal IgG antibodies have been shown in multiple studies to transfer to the infant *in utero*, there are little data to suggest that milk-derived antibodies are similarly transferred to the infant blood circulation during breastfeeding. To investigate whether passive immunity is conferred to the infant’s blood after maternal vaccination during lactation, we analyzed infant blood samples from mothers who were vaccinated after delivery (n=8). Blood samples were collected from these 8 infants (4 male, 4 female) at 68 days to 1 year of age (**Table S4**). Plasma was tested for the presence of anti-SARS-CoV-2 IgG and IgM and anti-RBD IgA. We evaluated infant blood samples collected at time frame of 4-10 weeks after 2nd dose as this time point corresponded to high anti-SARS-CoV-2 IgG levels in mothers’ blood and milk (**Figure 2 and 3**). No antibodies were detected in the blood of nursing infants born to mothers who were vaccinated postpartum (**Figure 5**), despite high IgG levels in maternal blood and milk. Infants born to mothers who received both doses of vaccine during pregnancy had detectable plasma anti-SARS-CoV-2 IgG levels at birth (15) and at follow-up (data not showed). None of the follow-up infant blood samples had detectable levels of anti-RBD IgA antibodies. These results demonstrate that vaccination during lactation induces anti-SARS-CoV-2 antibodies in human milk, but do not provides additional transfer of anti-SARS-CoV-2 antibodies to the infant blood, in contrast to vaccination during pregnancy.

**Figure 5:**
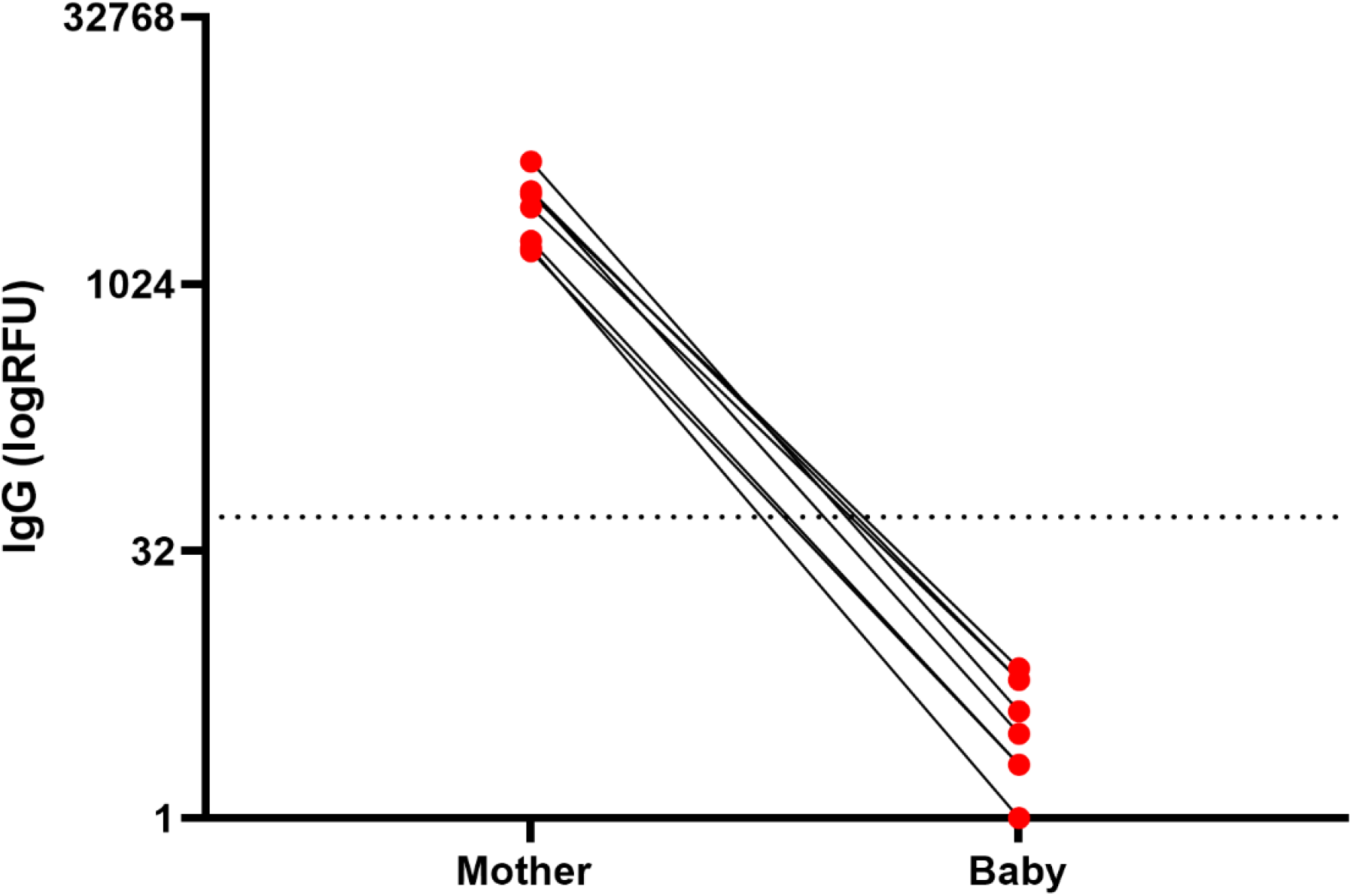
Infants anti-SARS-CoV2 IgG levels after maternal vaccination during lactation. IgG levels were measured in blood samples of infants and mothers 61 days to 1 year postpartum, 61-129 days after 1st maternal vaccine administration. Maternal and infant samples were collected in the same week (except in one case in which the maternal sample was collected 18 days prior to the infant sample). (RFU-relative fluorescent units, dashed line represents positive cut-off >50 RFU). Sample characteristics and individual antibodies levels are present in **Table S4**.

Lastly, we also investigated whether ingested antibodies could survive passage through the infant gastrointestinal tract by testing infant stool for anti-SARS-CoV-2 antibodies after vaccination (**Figure S2)** in one mother-infant dyad. We found that IgG (but not IgA) was detected in infant stool samples and were correlated with maternal milk anti-SARS-CoV-2 IgG levels.

## Conclusion

Our study provides a detailed report on patient symptoms and antibody responses of the COVID-19 mRNA vaccines in lactating mothers. We found that the rates of reported symptoms were similar to the CDC report from the V-Safe registry(16) but higher than described in the clinical trials, although we do not have a non-lactating comparison group. Importantly, there were no significant side effects noted in the infants of mothers vaccinated during breastfeeding. Comparing the mRNA vaccines made by current manufacturers, we found that lactating individuals may experience more vaccine-related side effects after mRNA-1237 compared to BNT-162b2 vaccine; however, no differences in immune response were observed between those vaccines.

It has recently been shown in a few studies that vaccine mRNA is not present in milk samples after vaccination(17,18), or is only detected in very low levels in some cases(19), providing reassurance that risks of exposure to the breastfed infant are minimal. This study adds to the evidence that vaccine components are minimally transferred to human milk. We found no significant increase in milk PEG-ylated protein concentrations at various time points after vaccine administration in a subset of samples analyzed in our cohort. We did observe one sample with higher ratio of PEG-ylated proteins 24 hours post vaccination (**Figure 1**, patient 030). This sample had PEG-ylated protein levels equivalent to 2.8µl/ml vaccine. However, we cannot confirm that the increased PEG in this single sample was from the COVID-19 vaccine, as PEG exposure may also be from other sources, such laxatives or ibuprofen. There was no increase in protein PEG-ylation ratio after 2nd dose in the same individual, and no unusual symptoms were reported in either the mother or her infant. Larger studies are needed to increase our understanding of transfer of PEG into human milk, and potential effects after ingestion by the infant. Although expert consensus states there is minimal or no potential risk for the infant from maternal COVID-19 vaccination(20,21), the minor symptoms that were reported (sleep changes and gastrointestinal symptoms) could be further investigated in future studies to determine if they are related to vaccination. Our findings also suggest that administration of maternal mRNA-based vaccine during lactation did not lead to a detectable immune response in the infant blood, which further suggests that the vaccine or its downstream products (e.g. spike protein) is not transferred to the infant via milk and cannot trigger infant immune responses via that route.

We also demonstrate that COVID-19 mRNA vaccination induces significant increases in anti-SARS-CoV-2 IgM and IgG levels in lactating mothers’ blood. Consistent with previous studies that showed IgM levels plateaued 28 days after COVID-19 infection (22), our results also demonstrated that dose 2 did not induce significantly higher levels of IgM than was is observed after dose 1. In contrast, maternal blood IgG levels increased by 6-fold after the 2nd dose (compare to the levels after the 1st dose), highlighting the importance of the 2nd dose to boost the antibody response (23).

Due to the lack of data about vaccination during pregnancy, many pregnant individuals were initially denied access to, declined, or were recommended to delay vaccination until after pregnancy. As such, many mothers have waited until after delivery to receive the vaccine. Although mothers vaccinated during lactation transferred antibodies to their infant through milk, which is an important component of mucosal immunity for the baby, there was no passive transfer of antibodies to the infant bloodstream, as occurs if the mother is vaccinated during pregnancy. Correlates of infant immune protection to COVID-19 are not yet well understood, however passive *in utero* transfer of IgG to the infant is important in the prevention of a number of infections including pertussis and influenza (24–26). Passively-transferred milk-derived IgA and IgG likely provide partial mucosal immune protection in infants, as breastfeeding is associated with lower risk of infections associated with mucosal defense, especially against respiratory infections (27–29).

Two nursing infants in our cohort were infected with COVID-19 during the study (one a week post maternal 2nd dose, and the second one between 1st and 2nd maternal vaccine), indicating that milk antibodies cannot fully protect against SARS-CoV-2 infection, especially at the time before full immune response is achieved in the vaccinated mother, typically 2-3 weeks after the booster dose (**Figure 2D**). Further studies are needed to determine the degree of protection conferred by IgA and IgG anti-SARS-CoV-2 antibodies that are present in milk. Interestingly, we demonstrated for the first time that anti-SARS-CoV-2 RBD IgG antibodies are detectable in stool samples collected 4 and 8 weeks post maternal vaccine (**Figure S2**), which suggests that milk-derived antibodies can persist in the infant gastrointestinal tract and provide protection against viral infection via the digestive system (30). Further studies evaluating the additive benefit of both transplacentally-derived maternal IgG, as well as milk-derived IgA and IgG are needed to determine protection against COVID-19 in early infancy. Our findings underscore the importance of determining the optimal timing of vaccine administration to confer maximal protection against COVID-19 in infancy.

Twenty-five percent of women in our cohort had no detectable levels of anti-RBD IgA in their milk after vaccination. Similar findings were reported in other studies (5,6), suggesting that production and transfer efficiency vary between individuals. Our analysis showed a negative correlation between infant age and milk anti-RBD IgA levels, which might explain some of the variation in milk IgA levels observed between different individuals. Ten out of the 12 participants who had no detectable anti-RBD IgA, had infants older than 5.5 months at the time of sample collection. The relationship between infant age, breastfeeding exclusivity, milk IgA antibodies, and optimal timing of vaccination during lactation remains to be studied in detail.

Strengths of our study include the prospective design and comprehensive symptom reporting by the vaccinated participants. We also report on longitudinal follow-up of infant immune responses, which has not been previously described. Furthermore, we included both BNT162b2 and mRNA-1273 vaccines and compared responses between the two vaccine manufacturers. Limitations include the small sample size, and that not all samples were able to be collected from all infant participants.

In summary, our study reports that no severe adverse events were noted in lactating individuals or their breastfeeding infants after COVID-19 mRNA vaccination. We demonstrated that human milk confers passive immunity to the infants, primarily through mucosal immunity in the gastrointestinal tract provided by IgA and IgG in milk. These results are important evidence to aid in counseling lactating individuals on the safety and efficacy of the COVID-19 mRNA vaccines, and the potential benefits to both the mother and infant.

## Data Availability

All data will be available

## Author contribution

YG designed the study, conducted experiments, acquired data, analyzed data, and was lead author of the manuscript. MP designed the study, conducted experiments, acquired data, analyzed data, assisted with editing and writing the manuscript. AC recruited participants, acquired data, and assisted with editing and writing the manuscript. CG analyzed data, conducted statistical analysis assisted with editing the manuscript. AHBW providing reagents and acquired data. UJ collected samples and conducted experiments. CYL recruited participants and collected samples. VJG, LW, SB, LL collected and process samples. EB collected and processed samples and assisted with editing the manuscript. IVA assisted with study and questionnaires design and editing and writing the manuscript. NA supervised the study and assisted in writing the manuscript. APM provided funding and assisted with editing the manuscript. SLG and VJF designed and supervised the study and assisted in writing the manuscript.

## Funding

These studies were supported by the Marino Family Foundation (to M.P), the National Institutes of Health (NIAID K23AI127886 to M.P. and NIAID K08AI141728 to S.L.G.), the Krzyzewski Family (to A.P.M. and S.L.G), the Weizmann Institute of Science - National Postdoctoral Award Program for Advancing Women in Science (to Y.G.), the International Society for Research In Human Milk and Lactation (ISRHML) Trainee Bridge Fund (to Y.G.), and of the Human Frontier Science Program (to Y.G.).

## Notes

**Conflict of interest:** The authors have declared that no conflict of interest exists.

### Competing Interest Statement

The authors have declared no competing interest.

### Author Declarations

The University of California San Francisco (UCSF) institutional review board approved the study (20-32077, 20-30410).

