## supplementary materials for "COVID-19 mRNA Vaccination in Lactation: Assessment of adverse events and vaccine related antibodies in mother-infant dyads"

**Supplementary data:**

**Methods:**

Infant stool samples were collected by the mothers to a sterile stool collection tube and were frozen immediately. Samples were kept in -80 until analysis. After thaw, infant stool samples were diluted 1:5 in stool preservative buffer (PBS, 0.05m EDTA, 1X proteinase inhibitor). Samples were later diluted 1:10 in sample dilutant buffer (Ray-Biotech, GA, USA, IEQ-CoVS1RBD-IgG-1 and IEQ-CoVS1RBD-IgA-1), filtered through a 0.2µm filter, and assayed by ELISA using the same protocol above used for milk.

**Results:**

***Anti SARS-CoV-2 RBD IgG antibodies were detected in infant stool samples:*** We further examined whether the anti SARS-CoV-2 IgG antibodies that were consumed by breastfeeding infants could survive the acidic conditions and enzymatic reactions in the intestinal tract. To address this question, we collected stool samples from one infant at 4 time points, pre-vaccine, on the day after the 2nd dose, and 4 and 8 weeks after the 2nd dose. We also collected paired samples of maternal milk at the same time points and measured these for the presence of anti SARS-CoV-2 IgG and IgA antibodies using ELISA. We were able to detect anti-SARS-CoV-2 IgG antibodies in post-vaccine samples collected 4 weeks and 8 weeks following the 2nd dose. These antibodies were not present in the pre-vaccine stool samples or in the sample collected one day after the 2nd dose (when milk IgG levels were still low) (**Figure S2).**

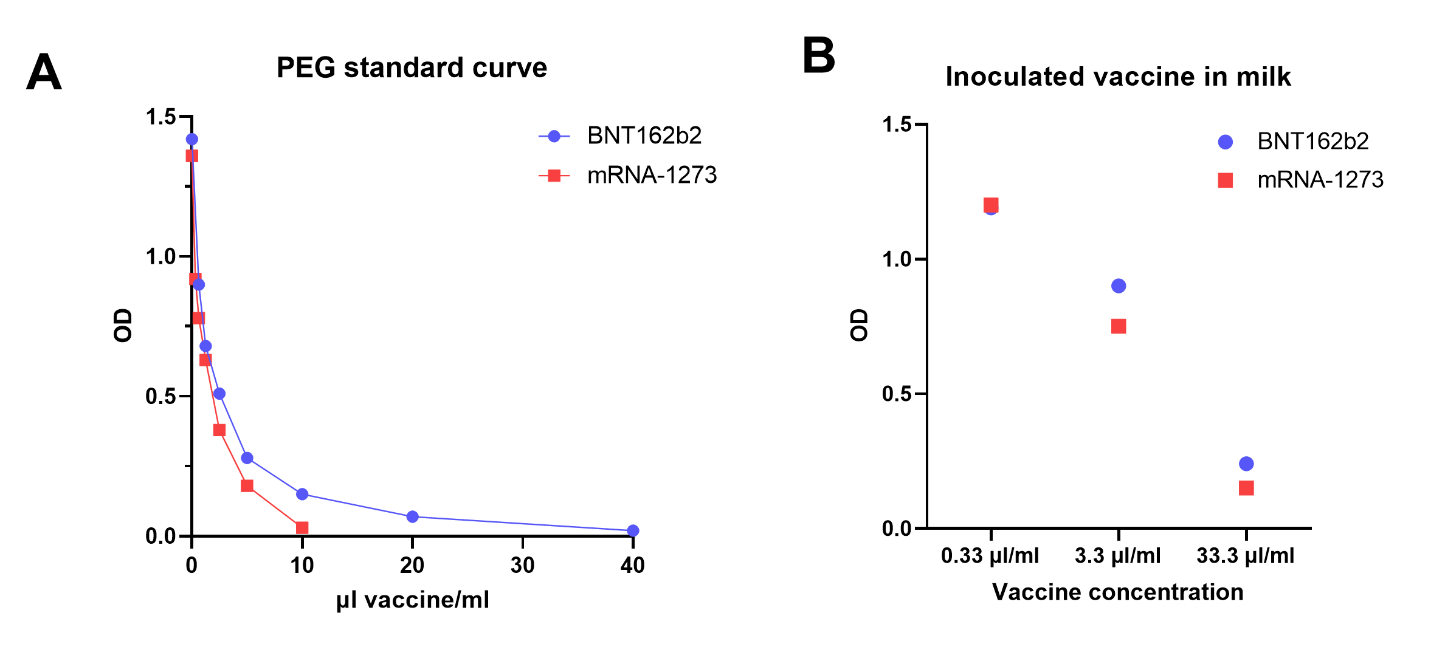

**Figure S1: Calibration of PEGylated protein assay.** A) different vaccine concentrations were used to generate a standard curve of OD (Y axis) vs. vaccine concentration (X axis). Vaccine concentrations in each sample were interpolated based on Sigmoidal, four-parameter logistic (4PL) curve. B) mRNA-1273 and BNT-162b2 vaccines were inoculated separately from pre-vaccine milk samples and were used to ensure the assay’s sensitivity to detect the vaccine PEG components in milk samples.

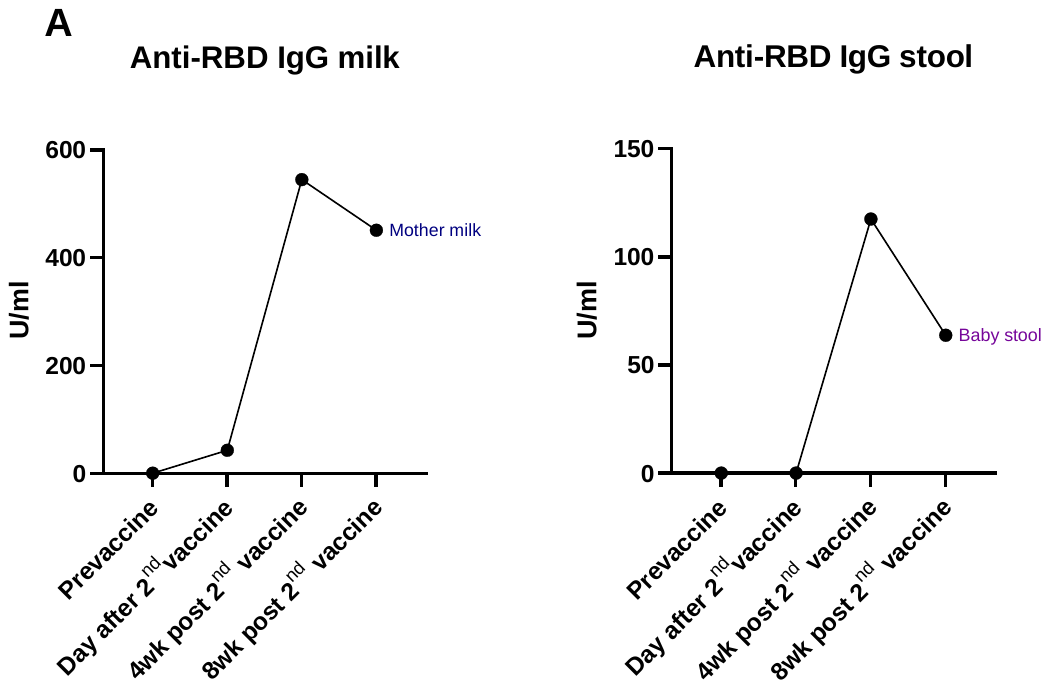
s

**Figure S2: Detection of** **anti-SARS-CoV2 RBD IgG in milk and stool samples.** Milk and stool samples were collected from one mother-baby dyad at various time points (X axis) and were assayed by ELISA. Note: milk samples are diluted in 1:2 ratio in assay samples buffer, and stool samples are diluted 1:5 in stool preservative buffer and 1:10 in assay samples buffer.

**Table S1. Participants testing positive for SARS-CoV-2 during the study period**

| **Participants testing positive for SARS-CoV-2** |  |  |  |
| --- | --- | --- | --- |
| **Participant ID:** | 1 | 2 | 3 |
| **Mother positive for SARS-CoV-2** | No | **Yes** | **Yes** |
| **Infant positive for SARS-CoV-2** | **Yes** | **Yes** | No |
| **Other people in the household diagnosed** | Yes | N/A | No |
| **Time of diagnosis** | 1 week after 2nd dose | N/A | 10 day before 1st dose |
| **Baby exclusively breastfed** | No | Yes | No |
| **Maternal blood IgG (RFU)** |  |  |  |
| On the day of 2nd dose | 244 | 5503 | N/A |
| 4 weeks after 2nd dose | 2558 | 5290 | N/A |
| **Infant blood IgG (RFU)** |  |  |  |
| 4 weeks after 2nd vaccine dose | N/A | 1928 | N/A |
| **Infant blood IgA (U/ml)** |  |  |  |
| 4 weeks after 2nd vaccine dose | N/A | 122 | N/A |
| **Milk anti-RBD IgG levels (U/ml)** |  |  |  |
| Pre-vaccine | N/A | 55 | 7.3 |
| On the day of 2nd dose | 246 | 2653 | 323 |
| 4 weeks after 2nd dose | 375 | 2834 | 250 |
| **Maternal blood positive for anti SARS-CoV2-N-protein antibodies** | N/A | Samples collected on day of 2nd dose and 5 weeks after 2nd dose were positive. | N/A |
| **Maternal injection site symptoms** |  |  |  |
| Reported after 1st vaccine dose: | Pain | Pain, Itching | None |
| Reported after 2nd vaccine dose: | Pain | None | Pain |
| **Maternal generalized symptoms** |  |  |  |
| Reported after 1st vaccine dose: | None | None | None |
| Reported after 2nd vaccine dose: | Fever, Chills, Muscle aches or body aches, Fatigue or tiredness | Fever, Chills, Fatigue or tiredness | Fatigue or tiredness |
| **Baby symptoms** |  |  |  |
| After 1st dose | None | None | None |
| after 2nd dose: | None | Less active. Feverish. | None |

**Table S2. Correlations between antibody levels and timing of samples in relation to childbirth and vaccine**

| Antibodies and sample types being correlated | N | Spearman correlations | |
| --- | --- | --- | --- |
|  |  | rho | p |
| Samples collected after 1st dose |  |  |  |
| IgG in maternal blood and … | 24 |  |  |
| Time from childbirth to collection |  | 0.38 | 0.07 |
| Time from 2nd dose to sample |  | -0.11 | 0.59 |
| IgG in breast milk and … | 35 |  |  |
| Time from childbirth to sample |  | -.05 | 0.75 |
| Time from 2nd dose to sample |  | -0.12 | 0.47 |
| IgA in breast milk and … | 38 |  |  |
| Time from childbirth to sample |  | -0.18 | 0.28 |
| Time from 2nd dose to sample |  | 0.12 | 0.45 |
| Samples collected after dose 2 |  |  |  |
| IgG in maternal blood and … | 32 |  |  |
| Time from childbirth to sample |  | 0.01 | 0.92 |
| Time from 2nd dose to sample |  | -0.34 | 0.05 |
| IgG in breast milk and … | 44 |  |  |
| Time from childbirth to sample |  | 0.04 | 0.75 |
| Time from 2nd dose to sample |  | 0.21 | 0.16 |
| IgA in breast milk and … | 43 |  |  |
| Time from childbirth to sample |  | **-0.35** | **0.02** |
| Time from 2nd dose to sample |  | -0.17 | 0.27 |

**Table S3. Antibody levels were not significantly correlated with maternal BMI**

| Correlation between maternal BMI and the following antibody levels: | n | Spearman correlations | |
| --- | --- | --- | --- |
|  |  | rho | p |
| After 1st dose |  |  |  |
| IgG in maternal blood | 24 | -.071 | .74 |
| IgG in milk | 35 | -.029 | .87 |
| IgA in milk | 38 | -.204 | .22 |
| 4-10 weeks after 2nd dose |  |  |  |
| IgG in maternal blood | 32 | .113 | .54 |
| IgG in milk | 44 | -.101 | .51 |
| IgA in milk | 43 | .115 | .46 |

**Table S4. Follow up maternal and infant blood samples characteristics and anti-SARS-CoV2 IgG levels**

| Participant # | Participant Cohort: | Infant sex | Infant age at sample date (months): | Infant age at 1st dose (months) | Time since 1st dose until infant sample collection (days) | Infant anti-SARS-CoV2 IgG antibodies (RFU): | Maternal anti-SARS-CoV2 IgG antibodies (RFU): | Vaccine type: |
| --- | --- | --- | --- | --- | --- | --- | --- | --- |
| 1 | Lactating | Female | 0-3 | 0-3 | 84 | 6 | 2768 | mRNA-1237 |
| 2 | Lactating | Male | 0-3 | 0-3 | 58 | 2 | 1791 | mRNA-1237 |
| 3 | Lactating | Female | 0-3 | 0-3 | 64 | 0 | 1638 | BNT162b2 |
| 4 | Lactating | Female | 3-6 | 0-3 | 67 | 4 | 5028 | mRNA-1237 |
| 5 | Lactating | Female | 3-6 | 0-3 | 61 | 6 | 3449 | mRNA-1237 |
| 6 | Lactating | Male | 6-9 | 3-6 | 81 | 7 | 3292 | mRNA-1237 |
| 7 | Lactating | Male | 9-12 | 6-9 | 103 | 2 | 1574 | mRNA-1237 |
| 8 | Lactating | Male | >12 | 9-12 | 62 | 3 | 3418 | BNT162b2 |

Note: negative values in infant age represent days before delivery. RFU>50 considered as positive value.
